# Ultra-Rare Genetic Variation in Relapsing Polychondritis: A Whole-Exome Sequencing Study

**DOI:** 10.1101/2023.04.10.23288250

**Authors:** Yiming Luo, Marcela A. Ferrada, Keith A. Sikora, Cameron Rankin, Hugh Alessi, Daniel L. Kastner, Zuoming Deng, Mengqi Zhang, Peter A. Merkel, Virginia B. Kraus, Andrew S. Allen, Peter C. Grayson

## Abstract

**Objective:** Relapsing polychondritis (RP) is a systemic inflammatory disease of unknown etiology. The study objective was to examine the contribution of rare genetic variations in RP.

**Methods:** We performed a case-control exome-wide rare variant association analysis including 66 unrelated European American RP cases and 2923 healthy controls. Gene-level collapsing analysis was performed using Firth’s logistics regression. Pathway analysis was performed on an exploratory basis with three different methods: Gene Set Enrichment Analysis (GSEA), sequence kernel association test (SKAT) and higher criticism test. Plasma DCBLD2 levels were measured in patients with RP and healthy controls using enzyme-linked immunosorbent assay (ELISA).

**Results:** In the collapsing analysis, RP was associated with higher burden of ultra-rare damaging variants in the *DCBLD2* gene (7.6% vs 0.1%, unadjusted odds ratio = 79.8, p = 2.93 x 10^-7^). Patients with RP and ultra-rare damaging variants in *DCBLD2* had a higher prevalence of cardiovascular manifestations. Plasma DCBLD2 protein levels were significantly higher in RP than healthy controls (5.9 vs 2.3, p < 0.001). Pathway analysis showed statistically significant enrichment of genes in the tumor necrosis factor (TNF) signaling pathway driven by rare damaging variants in *RELB*, *RELA* and *REL* using higher criticism test weighted by degree and eigenvector centrality.

**Conclusions:** This study identified specific rare variants in *DCBLD2* as putative genetic risk factors for RP. Genetic variation within the TNF pathway is also potentially associated with development of RP. These findings should be validated in additional patients with RP and supported by future functional experiments.

## Introduction

Relapsing polychondritis (RP) is a systemic inflammatory disease of unknown etiology that affects cartilage and other organs. RP is a rare disease with an estimated incidence of 3.5 cases per million (1). Patients with RP often have substantial diagnostic delay with significant tissue damage from uncontrolled inflammation. To date, there are no approved therapies for RP. Thus, there is an unmet need to understand casual factors and to identify potential therapeutic targets in RP.

Genetic association studies have played pivotal roles in understanding mechanisms of complex diseases and aid in discovery of effective therapeutics. Genetic association studies can be divided into common and rare variant association studies. Studying genetic risk from common variants often requires recruitment of large sample sizes, which is difficult in rare conditions like RP. Meanwhile, rare variant association studies with collapsing designs have potential to achieve substantial statistical power with a relatively smaller sample size (2).

The genetics of RP are poorly understood. Previous studies using targeted HLA genotyping have identified specific HLA associations with RP, including HLA-DRB1*16:02, HLA-DQB1*05:02, and HLA-B*67:01. Recently, somatic mutations in *UBA1* have been associated with VEXAS (vacuoles, E1 enzyme, X-linked, autoinflammatory, somatic) syndrome, a monogenic form of RP with a unique phenotype that includes chondritis, hematologic abnormalities, poor response to immunosuppressive therapy, and high mortality rate (3). Exome-wide genetic association studies have not been performed in RP. By focusing on coding regions, the functional impact of genetic variation is easier to predict compared to genetic variants present in non-coding regions. Familial clustering of RP has been reported (4), which suggests a possible role of germline genetic variation.

Thus, we performed a case-control genetic association study based on whole exome sequencing (WES) data in RP. The study objective was to identify genetic risk of rare coding variants with predicted deleterious effects in RP at the gene and pathway level.

## Materials and Methods

### Study Cohort

Patients with RP were enrolled into an ongoing prospective observational cohort study at the National Institutes of Health. Only patients who met the following criteria were included in this study: 1. Clinically diagnosed RP with objective evidence of chondritis defined as physician-observed erythema and swelling of the nose or ear consistent with nasal or auricular chondritis; upper airway inflammation confirmed by laryngoscopy; or damaging sequelae of chondritis such as cauliflower ear, saddle nose deformity, subglottic stenosis or tracheomalacia. 2. Self-reported ancestry being non-Hispanic European American. 3. WES data available by January 2021. There were no age restrictions. If two or more biologically related patients with RP were eligible for inclusion, we randomly included only one family member.

All patients underwent a standardized clinical and laboratory assessment, including a detailed medical history, physical exam, Physician Global Assessment (PhGA), audiology, otolaryngologist evaluation, dynamic computed tomography (CT) of the chest, and pulmonary function test (PFT). Patients who were treated with TNF inhibitors (TNFi) (adalimumab, etanercept, infliximab, golimumab and certolizumab) were assessed by the treating physician for therapeutic response based on clinical improvement of inflammatory manifestations. Patients who had improvement of their inflammatory manifestations at the next clinical visit after starting TNFi without the need to further increase steroid therapy were considered responders to TNFi. Patients were followed from January 2017 to June 2022.

Control individuals were included from the Atherosclerosis Risk in Communities (ARIC) cohort study (5). The ARIC cohort consists of individuals randomly sampled from their community (Forsyth County, NC; Jackson, MS; the northwest suburbs of Minneapolis, MN; and Washington County, MD) for cardiovascular preventive studies. The phenotype and genotype data of the ARIC cohort are publicly available at dbGaP (accession number: phs000280.v8.p2). 3,000 sex-matched, non-Hispanic European Americans with WES data were selected as controls for this study. Individuals with high coverage depth of WES data were prioritized for selection to minimize risk of having low-quality data.

### Whole Exome Sequencing and Bioinformatic Processing

WES was performed on peripheral leukocyte DNA from patients with RP using Agilent 51Mb Human Exome V5 (Agilent Technologies) capture and PE100-125 Illumina HiSeq2500 sequencing at Otogenetics Corporation (Georgia, U.S.) with a 100x average read coverage. For control individuals, WES was performed using NimbleGen SeqCap EZ VCRome (Roche, Basel, Switzerland) capture and Illumina HiSeq sequencing at Human Genome Sequencing Center at Baylor College of Medicine (Texas, U.S.) with a 92x average read coverage (6).

The raw sequencing data (fastq format) from patients with RP and controls were processed by the same bioinformatic pipeline following the best practice guideline by Genome Analysis Toolkit (GATK) (Broad Institute, Massachusetts, U.S.). Subsequent processing, including additional quality controls, coverage harmonization, principal component analysis (PCA) and Identity-by-descent analysis, were described in **Supplement Material**. A flowchart of the detailed bioinformatic and statistical methods is provided in **Supplemental Figure 1**.

### Sanger Sequencing

Custom primers were designed using Primer3, USCS Genome Browser, and NCBI BLAST for three regions of interest in the *DCBLD2* gene: Q435fs-F 5’-GGAATAAACTGACATCCGAGCA-3’ and Q435fs-R 5’-GCCCTGACCTCTATTGATTTGA-3’, I514F-F 5’-CTGCAACACAGTGGCCTTCTC-3’ and I514F-R 5’-CTCCTCGGAACAGCAATGACC-3’, and G261V-F and L250X-F 5’-TTCTTAGCCCAGTGCTACCC-3’ and G261V-R and L250X-R 5’-GAACTCCAGAGTAGTTAGAGGTGTG-3’. PCR was performed using a master mix of custom forward and reverse primers, OneTaq Hot Start 2x Master Mix with Standard Buffer from NEB, and water. Thermocycling conditions followed 95℃ for 3 minutes followed by 34 cycles of 94℃ for 30 secs, 60℃ for 30 secs, and 68℃ for 60 secs, ending with a final step of 68℃ for 5 minutes. PCR products were purified using Qiagen QIAquick PCR Purification Kit. PCR products were checked for site specificity by gel electrophoresis, and sequenced by ACGT DNA Sequencing Services (Germantown, MD, USA) to confirm the variant calling validity of the *DBCLD2* ultra-rare damaging QVs from WES.

The validation of Q435fs variant was performed after bacterial transformation. The PCR product was ligated into pGEM®-T Easy Vector (ProMega). Plasmids containing the PCR product were transformed into JM109 High Efficiency Competent E. coli using a heat-shock method (ProMega). Bacterial cells were plated on LB/IPTG/X-GAL/Amp plates and incubated overnight at 37°C. After overnight incubation, individual white colonies were transferred to bacterial culture tubes with LB/Amp liquid medium and incubated overnight at 37°C with shaking. Plasmids were then isolated from E. coli via ProMega miniprep kit specifications. After plasmid isolation, samples were sent to ACGT for sequencing using universal primers T7 and SP6.

### Enzyme-Linked Immunosorbent Assay (ELISA)

Plasma expression levels of DCBLD2 were quantified by ELISA (R&D Systems, Minneapolis, MN, USA) in 30 patients with RP (5 with *DCBLD2* ultra-rare QVs) and 32 healthy controls from the National Institutes of Health Blood Bank in accordance with the manufacturer’s instructions. RP patients with and without *DCBLD2* ultra-rare QVs were also selected for comparison. Patients were selected based on comparable PhGA and immunosuppressive regimens.

### Statistical Analysis

For the collapsing analysis, we constructed 5 models based on different qualifying variants (QVs) criteria (**Supplement Table 1**). Protein-truncating variants (PTVs) were annotated as “high confidence” without any flag by Loss-Of-Function Transcript Effect Estimator (LOFTEE) (7).

Firth’s logistic regression (8) was used to compare the burden of QV in each collapsing model between RP cases and controls, adjusting for sex and the first 3 principal components (PCs). Exome-wide significance level was set as 2.5 x 10^-6^, which represents Bonferroni correction for 20,000 genes. Study-wide significance level was set as 6.7 x 10^-7^, which represents Bonferroni correction for 20,000 genes x 4 models. Firth’s logistic regression does not rely on the asymptotic theories and thus is appropriate for rare variant analysis (8). Additionally, this test allows for covariate adjustment, which could potentially minimize bias and increase statistical power. The method and result of power calculation is provided in **Supplemental Material**.

Quantile-quantile plot (QQ-plot) was used to examine the distribution of observed p values against p values from a null distribution for each gene. The null distribution was obtained by permutation. In this process, the case and control labels were randomly permutated 1,000 times, and rank-order p values were obtained for each gene in each permutation set using Firth’s logistic regression. The expected p value in the null model for each gene was the mean of each rank-ordered p values across the 1,000 permutations. QQ-plot is a method to examine whether there is inflation of results from systematic bias, such as confounding due to population genetic structure.

We also performed exploratory analysis to examine whether RP was associated with rare genetic variants at the pathway level. Three pathway-analysis approaches were used including Gene Set Enrichment Analysis (“GEEA preranked”) (9), Sequence kernel association test (“SKAT_robust”) (10), and higher criticism test (11). The input of GSEA and higher criticism test was from genes ranked by p values from the gene-level collapsing analysis. For the higher criticism test, both unweighted and weighted higher criticism tests were performed. The weight was based on a gene intolerance index that was based on the Residual Variation Intolerance Score (RVIS) (12), degree centrality (13), eigenvector centrality (14) and RankPage centrality (15). The centrality calculation was based on gene interaction data from the bioGRID database (version 3.4.147) (16). Protein interaction networks were generated from the bioGRID database for top ranked genes in significant pathways from weighted higher criticism test.

The gene sets studied included all 50 gene sets of the “Hallmark Gene Sets” from the Molecular Signatures Database (MSigDB) (17). An additional three gene sets related to cartilage (“GOBP_REGULATION_OF_CARTILAGE_DEVELOPMENT”, “GOBP_CARTILAGE_DEVELOPMENT” and “HP_ABNORMAL_CARTILAGE_MORPHOLOGY”) were included in the analysis.

Test-level multiple comparison adjustment was applied for each collapsing model in each method in the pathway-level analysis. For GSEA, family-wise error rate (FWER) provided by the GSEA software (18) was used to adjust for multiplicity. For SKAT, Bonferroni correction for 53 gene sets was used to adjust for multiplicity. For the higher criticism test, the minP algorithm (19) was used to obtain multiplicity adjusted p values across all the gene sets analyzed. R (version 4.1.2) was used for all statistical analyses, including R packages “logistf”, “QQperm”, “SKAT” and “wHC”. This study was approved by the National institutes of Health Intramural Research Program Institutional Review Board (IRB Number: 14AR0200).

## Results

### Study Cohorts

There were 89 cases of RP with WES data available for this study. Among them, there were 5 pairs with parent-offspring relationship, and one of each pair was randomly excluded. Sixteen additional cases were excluded due to self-identified ancestry not being non-Hispanic European American. After additional quality controls to remove PCA outliers and cryptic relatedness, a total of 2,989 samples including 66 RP cases and 2,923 controls were included for association analysis (**Figure 1**). On average, 90.4% of the exonic sites were with well and balanced coverage and included. The percentage of exonic sites with well and balanced coverage in each gene are shown in **Supplemental Table 2**. The PCA plot for examining population genetic structure and ancestral clustering is shown in **Supplemental Figure 2**.

**Figure 1.**
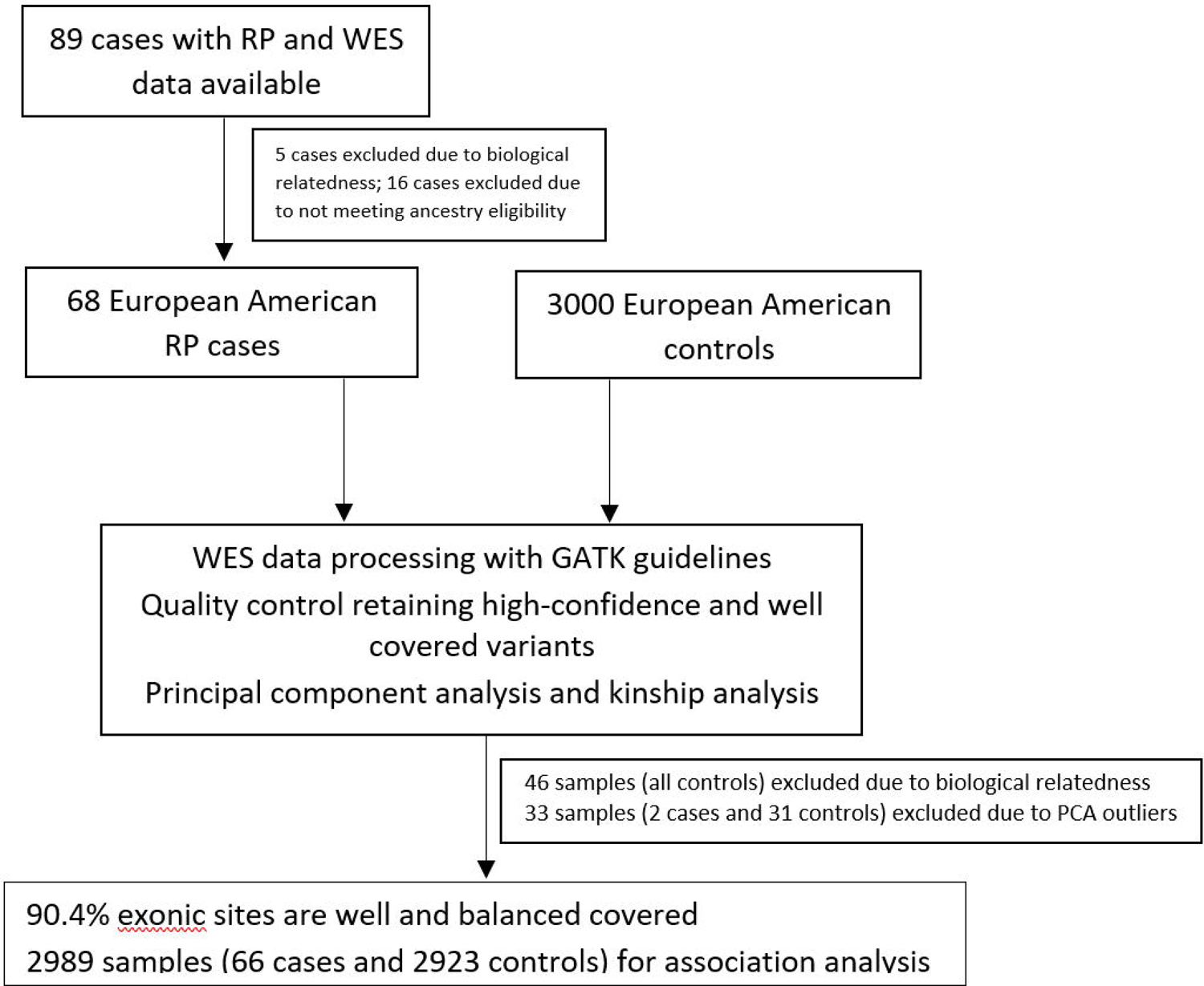
Flowchart of study population. 66 cases of relapsing polychondritis and 2923 healthy controls were ultimately included in this exome-wide rare variant genetic association study.

The clinical characteristics of the included RP cases are summarized in **Table 1**. Among the 66 included cases of RP, 77% were female sex. Mean age at symptom onset was 32 years, and mean age at diagnosis was 38 years. Auricular chondritis (64%), nasal chondritis (76%), costochondritis (42%), ocular inflammation (24%) and cutaneous inflammation (24%) were prevalent. Use of prednisone at doses ≥60mg daily (52% of patients) and treatment with biologic or targeted synthetic immunosuppressants (65% of patients) was common. Over the course of disease, 9% had been hospitalized in an intensive care unit, and 3% died over the course of this study.

**Table 1.** Clinical Features of 66 RP cases included for rare variant association analysis

|  |  |
| --- | --- |
| *Demographic Features* |  |
| Female sex | 77% |
| Age at the onset of symptoms (years, mean +- SD) | 32 +- 16 |
| Age at diagnosis (years, mean +- SD) | 39 +- 18 |
| *Clinical Manifestations<sup>a</sup>* |  |
| Auricular chondritis | 64% |
| Nasal chondritis | 76% |
| Airway chondritis | 42% |
| Costochondritis | 71% |
| Ocular inflammation | 24% |
| Cutaneous inflammation | 24% |
| Arthritis or arthralgia | 82% |
| Oral ulcerations | 24% |
| Genital ulcerations | 17% |
| Venous thromboembolism<sup>b</sup> | 5% |
| Raynaud phenomenon | 17% |
| CNS Involvement | 0% |
| Myocarditis | 0% |
| Pericarditis | 2% |
| Aortitis | 3% |
| ANA positive | 16% |
| *Treatment* |  |
| High-dose steroid<sup>c</sup> | 53% |
| Steroid-sparing immunosuppressants | 77% |
| Biological or targeted immunosuppressants | 65% |
| *Complications* |  |
| Death | 3% |
| Intensive Care Unit admission | 9% |
| Tracheotomy | 5% |
| Subglottic stenosis | 14% |
| Tracheomalacia | 38% |
| Bronchomalacia | 17% |

CNS: central nervous system; ANA: antinuclear antibody
- The clinical manifestations were recorded if they were ever present during the course of their illness. - Venous thromboembolism includes unprovoked deep venous thrombosis and pulmonary embolism. - High-dose steroid is defined as use of $\geq 60$ mg daily prednisone or steroid equivalent.

### Gene-Level Analysis

In the ultra-rare damaging model, only the *DCBLD2* gene reached both exome-wide and study-wide statistical significance (p = 2.93 x 10^-7^). Five of out 66 RP cases and 3 out of 2923 controls carried a *DCBLD2* QV in the ultra-rare damaging model (7.6% vs 0.1%, unadjusted odds ratio = 79.8). The QQ plot showed no evidence of bias due to the population genetic structure (**Figure 2**). *DCBLD*2 remained the top ranked gene for the rare damaging model (7.6% vs 0.5%, p = 3.91 x 10^-5^) and the rare PTV model (3% vs 0, p = 5.59 x 10^-5^). The sites of ultra-rare damaging QVs within the *DBCLD2* gene in patients with RP were located in regions of acceptable coverage depth in GnomAD (**Supplemental Table 3**). The amino acid position changes of the *DCBLD2* ultra-rare QVs and the affected domains are shown in **Supplemental Figure 3**.

**Figure 2.**
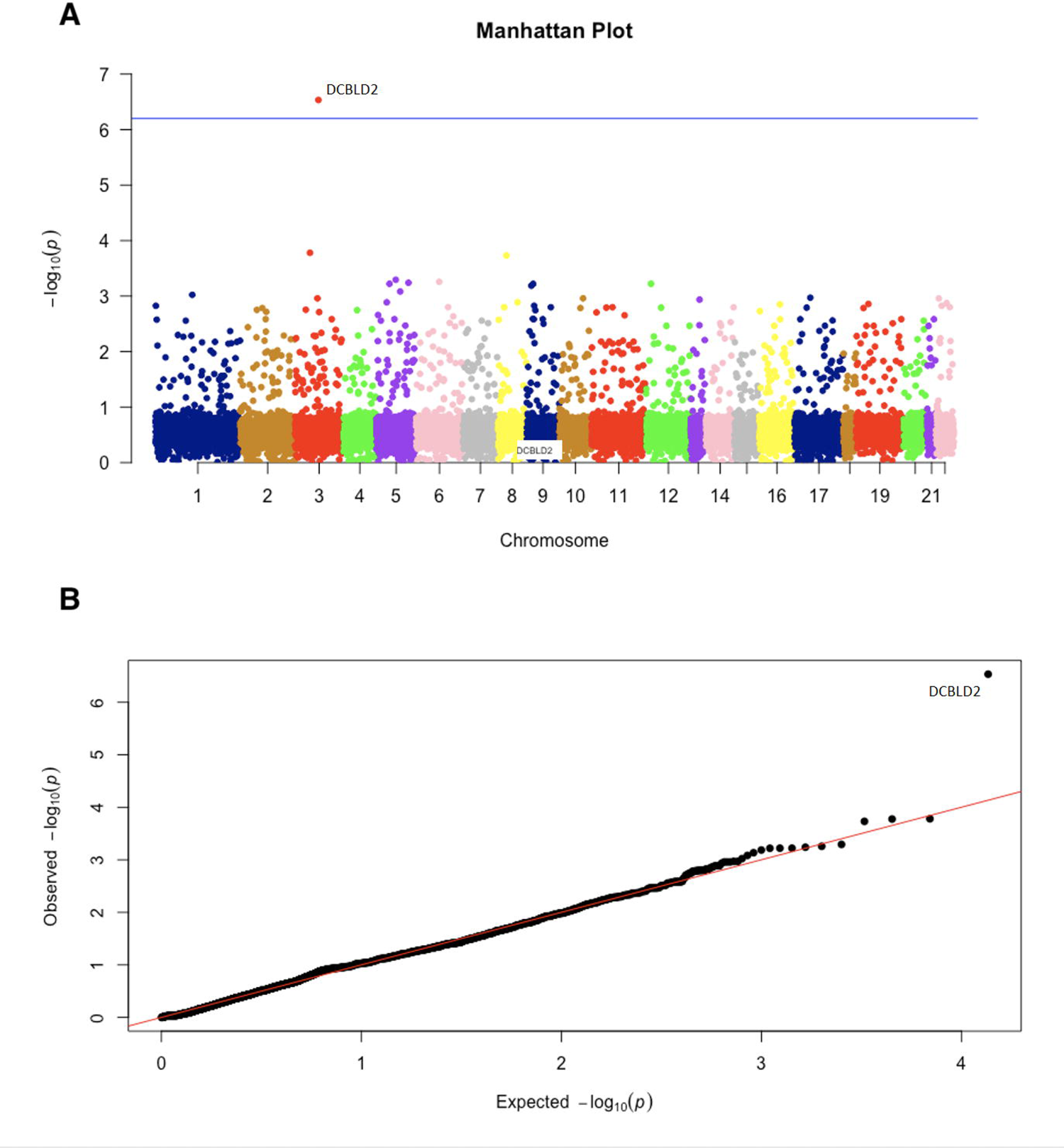
Manhattan plot and QQ plot of genetic associations for the ultra-rare damaging model. *DCBLD2* was associated with RP (7.6% vs 0.1%, unadjusted odds ratio = 79.8, p = 2.93 x 10^-7^) in the ultra-rare damaging model (primary model). QQ plot shows that the distribution of observed p values was consistent with permutation-based null distribution except for*DCBLD2*. This indicates that there is no evidence of systematic confounding bias due to population genetic structures

Rare somatic variants in *UBA1* were previously demonstrated to be pathogenic for VEXAS syndrome, a monogenic disease with a large degree of clinical overlap with RP (3). In this study, rare germline variants in *UBA1* were not increased in patients with RP versus controls (0/66 vs 4/2923 for the ultra-rare damaging model, 0/66 vs 23/2923 for the rare damaging model, 0/66 vs 0/2923 for the PTV model and 0/66 vs 1/2923 for the recessive model).

For the remaining collapsing models, no genes reached exome-wide or study-wide statistical significance (**Supplemental Figure 4, Supplemental Table 4**). Ultra-rare *DCBLD2* QVs were not found in the 16 non-European American patients with RP who were excluded from the association analysis due to not meeting ancestry eligibility.

### Familial Clustering and Clinical Phenotype of Patients with RP with *DCBLD2* Ultra-Rare QVs

At the variant level, there were 4 specific *DCBLD2* ultra-rare QVs identified among patients with RP: two QVs were missense variants (G261V and I514F); one QV was a stop-gain variant (L250X); and one QV was a frameshift variant (Q435fs). We next examined the*DCBLD2* ultra-rare damaging QVs in all 89 RP patients with WES available. Two additional RP patients, both first-degree relatives of patients with *DCBLD2* ultra-rare damaging QVs (G261V and Q435fs), had the same *DCBLD2* variant as their familial counterpart. For one RP patient who carried an ultra-rare damaging *DCBLD2* QV (L250X), the WES data for two first-degree healthy relatives were also available, and both had the same *DCBLD2* variant without clinical symptoms suggestive of RP. Thus, there were 7 patients with RP, among a total of 89 patients, with *DCBLD2* ultra-rare damaging QVs, as summarized in **Table 2**. The Pedigree plot of the three families with carriers of *DCBLD2* ultra-rare damaging QVs is shown in **Figure 3**. Sanger sequencing confirmed the validity of all the 4 discovered *DCBLD2* ultra-rare QVs in the 7 patients (**Supplemental Figure 5**).

**Figure 3.**
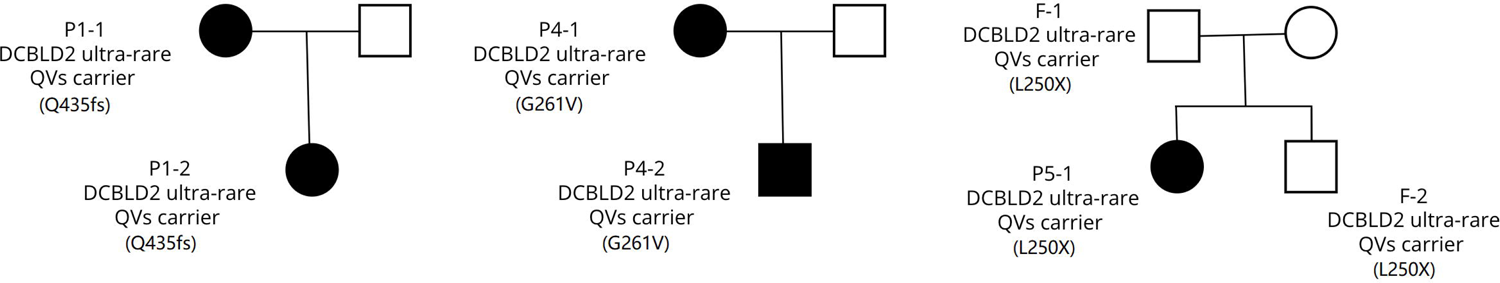
Pedigree plot of RP patients with DCBLD2 ultra-rare damaging QVs and genotype data from family members available. In the 5 unrelated patients with RP and DCBLD2 ultra-rare damaging QVs, 3 of them have genotype data of family members available. In two families, there is another first-degree family member who has RP and carries the DCBLD2 ultra-rare damaging QV. In the third family, there are two first-degree family members who carry the DCBLD2 ultra-rare damaging QV but do not have RP.

**Table 2.** DCBLD2 ultra-rare damaging QVs identified in patients with RP

| Variant position | Transcript affected | Impact | CADD score | Individuals affected |
| --- | --- | --- | --- | --- |
| 3:98531235-TG-T | DCBLD2:NM_080927:exon 10:c.1303delC;p.Q435fs | frameshift_deletion | 33 | 2 RP cases, mother and daughter |
| 3:98530074-T-A | DCBLD2:NM_080927:exon 12:c.A1540T;p.I514F | nonsynonymous_SNV | 24.4 | 2 RP cases, unrelated |
| 3:98541120-C-A | DCBLD2:NM_080927:exon 6:c.G782T;p.G261V | nonsynonymous_SNV | 28.2 | 2 RP cases, mother and daughter |
| 3:98541153-A-T | DCBLD2:NM_080927:exon 6:c.T749A;p.L250X | stopgain | 40 | 1 RP case, 3 healthy relatives of this patient |

Among the 89 cases of RP, the clinical characteristics of patients with and without*DCBLD2* ultra-rare damaging QVs are summarized in **Supplemental Table 5**. Compared to patients without *DCBLD2* ultra-rare damaging QVs, there was a numerically higher proportion of patients carrying *DCBLD2* ultra-rare damaging QVs with cardiovascular manifestations of disease, including venous thromboembolism (14% vs 5%), Raynaud’s phenomenon (29% vs 15%), pericarditis (14% vs 2%) and aortitis (14% vs 1%); however, statistical significance was not reached. Regarding additional cardiac manifestations in RP with *DCBLD2* ultra-rare damaging QVs, one patient had restrictive cardiomyopathy, and another patient had severe valvular heart disease in both mitral and tricuspid values and required mitral valvuloplasty. The clinical manifestations of patients with RP and *DCBLD2* ultra-rare damaging QVs are summarized in **Supplemental Table 6**.

### Pathway-Level Analysis

As expected, pathways containing *DCBLD2* genes, including “HALLMARK_APICAL_SURFACE”, “HALLMARK_INFLAMMATORY_RESPONSE” and “HALLMARK_KRAS_SIGNALING_UP”, were statistically significant or top ranked with multiple collapsing models across the 3 statistical methods. However, after removing the *DCBLD2* gene, these pathways were no longer significantly associated with a diagnosis of RP.

After removing the *DCBLC2* gene, “HALLMARK_TNFA_SIGNALING_VIA_NFKB” (not containing DCBLD2) was top ranked by both GSEA and unweighted higher criticism test in the rare damaging model, but did not reach multiplicity-adjusted statistical significance. With the weighted higher criticism test, “HALLMARK_TNFA_SIGNALING_VIA_NFKB” reached test-level statistical significance when weighted by degree centrality and eigenvector centrality, in both the ultra-rare damaging model (p = 0.039 weighted by degree centrality, p < 0.001 weighted by eigenvector centrality) and the rare damaging model (p = 0.039 weighted by degree centrality, p < 0.001 weighted by eigenvector centrality). The genes that were favorably weighted by degree or eigenvector centrality (defined as the weighted p values being smaller than the unweighted p values) and with at least one QV in RP in the two models were: *RELB, RELA, REL, ABCA1, NFE2L2, TRAF1, BIRC2, TNIP1, NR4A1, TNFAIP3* (**Supplemental Table 7**). After removing the top three genes RELB, RELA and REL, statistical significance was lost (p = 1 weighted by degree centrality and eigenvector centrality in both the ultra-rare damaging model and rare damaging model) (**Supplemental Table 8**). The protein interaction network of RELB, RELA and REL is shown in **Supplemental Figure 6-8**.

The results of the pathway analysis with each method are included in **Supplemental Tables 8-10**. The top ranked 3 pathways for each collapsing model with the 3 statistical methods are summarized in **Supplemental Table 11**.

### DCBLD2 Plasma Protein Levels

DCBLD2 plasma protein levels were measured using ELISA in 30 patients with RP (5 with *DCBLD2* ultra-rare QVs) and 32 healthy controls (HC). A total of 9 of 30 RP patients (30%) and 5 of 32 HC (16%) had plasma DCBLD2 levels above the assay detection range. After replacing all the values that were above the assay detection range with the highest level of quantification, plasma DCBLD2 levels were significantly higher in RP than HC (5.9 vs 2.3, p < 0.001). DCBLD2 levels were even higher in RP patients without *DCBLD2* ultra-rare QVs (6.6 vs 2.3 in HC, p < 0.001). In RP patients with *DCBLD2* ultra-rare QVs, the DCBLD2 levels were lower but still numerically higher than those in HC (2.6 vs 2.3, p = 0.075) (**Figure 4**).

**Figure 4.**
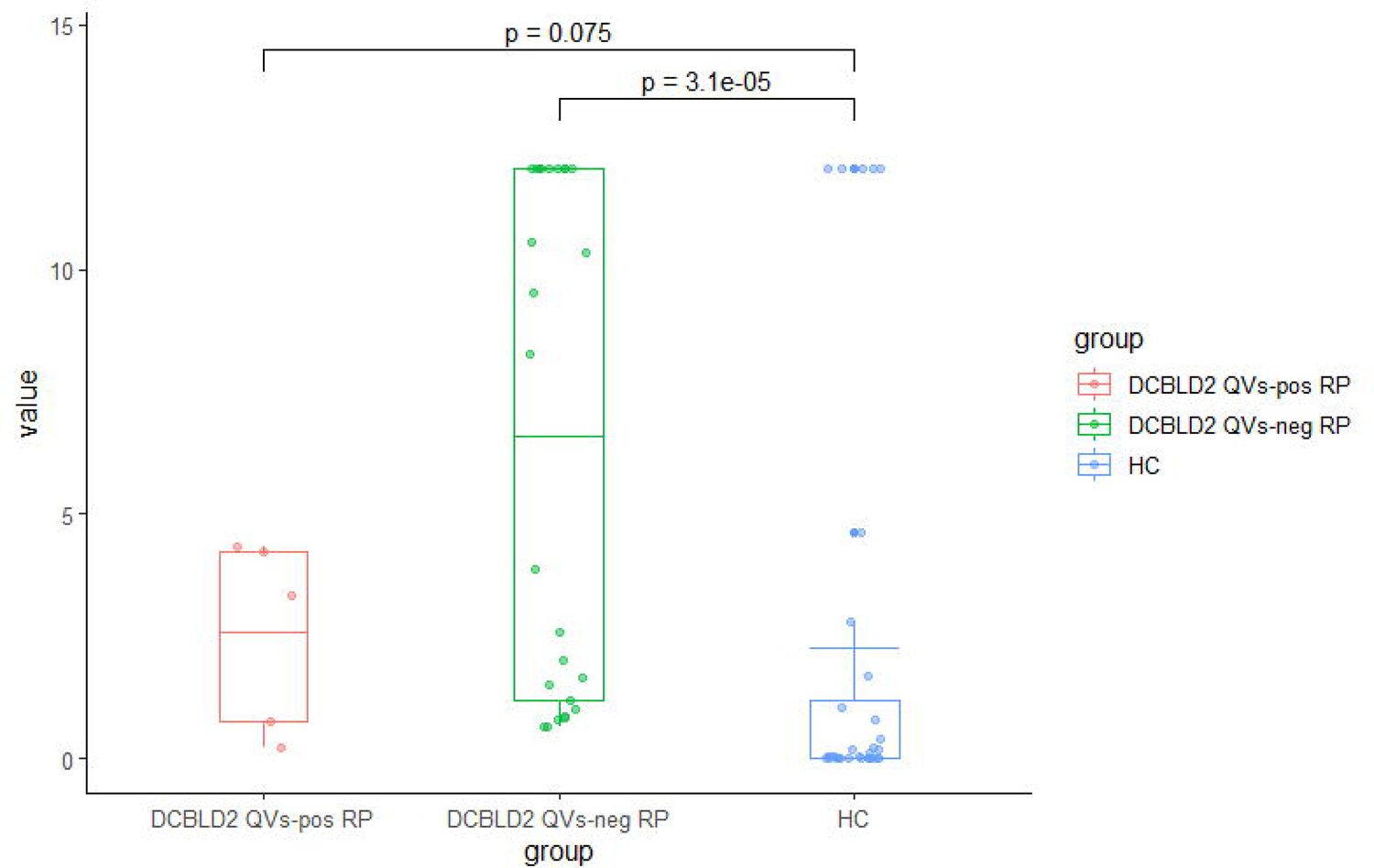
Plasma DCBLD2 protein levels in patients with RP and healthy controls. DCBLD2 levels were significantly higher in patients with RP but without DCBLD2 ultra-rare QVs compared to those in HC (6.6 vs 2.3 in HC, p < 0.001). In patients with RP and DCBLD2 ultra-rare QVs, the DCBLD2 levels were lower but still numerically higher than those in HC (2.6 vs 2.3, p = 0.075)

### Therapeutic Response to TNFi

Among the total 89 patients with RP, 48 (54%) were treated with TNFi. Six (13%) patients had at least one rare damaging QV in the “HALLMARK_TNFA_SIGNALING_VIA_NFKB” pathway (“TNF pathway”). One patient with a rare damaging QV in the TNF pathway who was treated with a TNFi was excluded due to loss of follow up before therapeutic response could be assessed. For the remaining 47 patients who were treated with a TNFi, all 5 RP (100%) patients with a TNF pathway rare damaging QVs and 23 of 42 (55%) RP patients without a TNF pathway rare damaging QVs responded to TNFi treatment (p = 0.072).

## Discussion

This is the first exome-wide rare variant association study performed in patients with RP. The results demonstrate that ultra-rare damaging variants in the *DCBLD2* gene are associated with a clinical diagnosis of RP. Further, examination of the *DCBLD2* ultra-rare damaging QVs in multiplex cases of RP revealed familial clustering in two mother-daughter pairs with RP, which provides additional evidence supporting an association between genetic variants within *DCBLD2* and RP. Taken together, *DBCLD2* is the first putative non-HLA gene with germline genetic variation contributing to the heritability of RP.

The full name of *DCBLD2* is Discoidin, CUB And LCCL Domain Containing 2. It is also called Endothelial And Smooth Muscle Cell-Derived Neuropilin-Like Protein (*ESDN*). DCBLD2 is a type I transmembrane protein that is primarily located in the plasma membrane (20). The extracellular region of DCBLD2 contains a signal sequence, followed by the CUB, LCCL, and Coagulation Factor V/VIII type-C (Discoidin) domains. All the *DCBLD2* ultra-rare damaging QVs identified in cases of RP were located in the extracellular region. The variant L250X and G261V are in the LCCL domain. The variant Q435fs is in the FV/FVIII domain, and the variant I514F is in the extracellular region close to the transmembrane region.

It is unlikely that genetic variants in *DCBLD2* cause a monogenic form of RP. Rather, genetic variation in *DCBLD2* is likely a risk factor that contributes to a complex disease process. The DCBLD2 ultra-rare damaging QVs contain both missense and PTV variants, which suggests that the functional impact of these variants is more likely loss of function (LoF). The “probability of being LoF intolerant score” (pLI score) of the *DCBLD2* gene is 0.1, which argues against its ability to cause haploinsufficiency with heterozygous LoF variation (21). The pLI score was developed as a dichotomous metric. A continuous metric named loss-of-function observed/expected upper bound fraction (LOEUF) was later developed to assess each gene along a continuous spectrum of tolerance to inactivation (7). The LOEUF for *DCBLD2* is 0.446, which suggests moderate evolutionary constraint against LoF variants. Additionally, the segregation pattern observed in this study in family members of patients with RP who have ultra-rare damaging QVs in *DCBLD2* also argues against Mendelian inheritance. In our dataset, two healthy relatives had the same ultra-rare damaging QVs in *DCLBD2* as their proband RP case. However, given the large statistical effect size, familial clustering and dramatically elevated DCBLD2 plasma protein levels observed in this study, the likelihood that the association between rare variants in *DCBLD2* and RP was merely by chance seems extremely low.

The mechanistic role of *DCBLD2* in RP is elusive. Our study observed a numerically higher proportion of cardiovascular manifestations in patients with *DCBLD2* ultra-rare damaging QVs, but the sample size was too small to definitively conclude that these ultra-rare variants lead to a distinctive phenotype of RP. Animal and *in-vitro* studies have suggested that *DCBLD2* is involved in cardiac function, vascular repair, and thrombosis (22–26). Recently, a child with a homozygous *DCBLD2* stop-gained variant (W27*) was reported (27). This patient had severe restrictive cardiomyopathy, neurovascular malformation, skeletal abnormalities, and died at the age of 5. However, no autoimmune or inflammatory features, including chondritis, were reported, and the functional implications of *DCBLD2* loss-of-function were not investigated.

Another way of interpreting this genetic association study is that it may inform a common causal mechanism of RP whereby a critical biological pathway involving *DCBLD2* is disrupted. In this study, significantly elevated plasma DCBLD2 protein levels, including those without*DCBLD2* ultra-rare damaging QVs, supports this hypothesis. Based on the information from the Human Protein Atlas (28, 29), there is low tissue specificity regarding *DCBLD2* expression. *DCBLD2* has low expression in immune cells and moderate expression in chondrocytes. As *DCBLD2* is known to be involved in vascular damage repair (22), it is possible that plasma DCBLD2 levels in RP are elevated secondary to tissue damage and that lower levels of *DCBLD2* in patients with ultra-rare damaging variants may render certain tissues, such as cartilage, more prone to inflammatory insult. A recent analysis revealed that upregulation of *DCBLD2* is associated with tumor immunosuppression across multiple cancer types (30).

Pathway analysis using three different analytic approaches demonstrated that genetic variation in the TNF signaling pathway is associated with a diagnosis of RP. This association was largely driven by rare genetic variants in three genes: *RELB*, *RELA* and *REL*. Pathway analysis to evaluate rare genetic variant association is exploratory in nature; a gold standard to examine the validity of the finding is lacking. RELB, RELA and REL are all members of the second class of Rel/NF-κB transcription factors that are involved in a variety of human immune and inflammatory responses (31). This study reported that 60% patients with RP responded to TNFi treatment, which is consistent with the efficacy results from a recent systematic review in RP (32). This study also found a numerically higher proportion of TNFi response in patients with RP who carry rare-damaging QVs in the TNF pathway. These clinical observations support the hypothesis that TNF pathway dysregulation plays a pivotal role in the pathogenesis of RP. The potential role of genetic sequencing in treatment selection warrants further investigation. This study also highlights the advantage of incorporating external information for signal detection in pathway-level rare variant association studies to identify potential therapeutic targets in rare diseases.

There are several limitations of this study. Foremost, the findings were not validated in an independent cohort, which due to the rarity of this disease, is challenging to recruit. Given the relatively small sample size, this study was primarily designed to evaluate ultra-rare variants with very large effect size and may not identify rare variants with smaller effects. The analysis was restricted to cases of RP with non-Hispanic European ancestry because there were too few cases of RP in the cohort from other ancestries for subsequent analysis. Although RP patients with rare variants in *DCBLD2* had numerically higher prevalence of cardiovascular manifestations, the sample size was too small to have sufficient statistical power to confirm these associations. The biological plausibility of an association between rare variants in*DCBLD2* and RP pathogenesis remains unclear. The cases and controls in our study were genotyped using different sequencing platforms, which can introduce false discovery. However, the proportion of *DCBLD2* ultra-rare variants in RP greatly exceeds the expectation of being extremely rare given our model design. In conclusion, in this exome-wide rare variant association study, we identified rare variants in *DCBLD2* and *TNF* pathway as putative genetic risk factors for RP.

## Data Availability

All data produced in the present study are available upon reasonable request to the authors

## Acknowledgement

None

