## Supplementary material for "Ultra-Rare Genetic Variation in Relapsing Polychondritis: A Whole-Exome Sequencing Study": Supplemental Material.docx

**Bioinformatic Processing**

The raw sequencing data (fastq format) from RP and controls were processed by the same bioinformatic pipeline following the best practice guideline by Genome Analysis Toolkit (GATK) (Broad Institute, Massachusetts, U.S.). The variants were annotated with ANNOVAR [1] and Ensembl Variant Effect Predictor (VEP) [2].

Additional quality control steps were performed with variants included only if they met the following criteria: (1) at least 10-fold coverage (2) QUAL at least 50 (3) GQ at least 20 (4) QD at least 2 (5) MQ at least 40 (6) RPRS greater than -3 (7) MQRS greater than -10 (8) FS <= 200 for indels and <= 60 for SNVs (9) VQSR tranche of 99% (10) Hardy–Weinberg equilibrium p value >= 0.001 (11) het alt allele ratio 0.3 to 0.7, ref-ref alt allele ratio < 0.15, hom alt allele ratio > 0.85 (12) genotype call rate >= 90% in cases and controls, respectively. Most of these criteria were based on a previously published exome-wide rare variant association study [3].

The coverage of each Consensus Coding Sequence (CCDS) site was obtained for all samples by Samtools [4]. For each CCDS site, the absolute difference of the percentage of samples with coverage ≥ 10 between RP and controls was calculated. A cut-off of the absolute difference was selected when the following conditions were met: (1) [(absolute difference cut-off) + (percentage of sites pruned)] was minimized; (2) absolute difference cut-off ≥ 5%; (3) percentage of sites retained ≥ 90%.

After the above quality control steps, principal component analysis (PCA) was performed using Eigensoft [5] over a subset of uncorrelated (r2 < 0.1) polymorphic markers from the Illumina HumanCore chip. This marker selection was consistent with a previously published study [6]. PC outliers were removed if they were away from the center and exceeded 6 standard deviations (SD), which is the default setting for Eigensoft.

Identity-by-descent analysis was performed using KING [7] inside Plink 2 [8] with the same set of polymorphic markers. One member of each pair was removed if the KING kinship coefficient was below 0.094.

**Power Calculation**

Statistical power was calculated using simulation with different levels of odds ratio (OR) tested given our sample size of 66 cases and 2923 controls. The probability of an individual carrying a QV is p1 for RP and p2 for controls, both assumed to follow a binomial distribution. We assumed p2=0.001. p1 will be calculated based on different OR tested. Firth’s logistic regression was performed 5,000 times with each simulation for each tested OR. The proportion of p values below 2.0 x 10^-6^ and 6.7x10^-7^ is the exome-wide and study-wide statistical power for each tested OR, respectively.

The result of power calculation with different levels of OR was as following:

OR=50, power(exome)=0.36, power(study)=0.30

OR=60, power(exome)=0.48, power(study)=0.42

OR=70, power(exome)=0.59, power(study)=0.54

OR=80, power(exome)=0.69, power(study)=0.65
 OR=90, power(exome)=0.78, power(study)=0.74

OR=100, power(exome)=0.84, power(study)=0.80

**Supplemental Table 1** Qualifying variants selection criteria for collapsing analysis

| Model names | VAF in GnomAD | Predicted deleterious effects | Others |
| --- | --- | --- | --- |
| Ultra-rare model (primary) | < 0.1% | non-synonymous, CADD >= 20 or PTV |  |
| Rare damaging model | < 0.1% | non-synonymous, CADD >= 20 or PTV |  |
| Rare PTV model | < 0.1% | non-synonymous, PTV |  |
| Recessive model | < 1% | non-synonymous, CADD >= 20 or PTV | at least biallelic |
| Synonymous model (negative control) | < 0.1% | synonymous |  |

VAF: variant allele frequency; GnomAD: the Genome Aggregation Database; CADD: Combined Annotation Dependent Depletion; PTV: protein truncating variant

**Supplemental Table 2** Percentage of exonic sites with well and balanced coverage in each gene

(Table in a separate file)

**Supplemental Table 3** DCBLD2 ultra-rare damaging QVs site mean coverage in GnomAD

| Variant position | Mean exome sample coverage | Percentage of exome samples with at least 10x coverage | Mean genome sample coverage | Percentage of genome samples with at least 10x coverage |
| --- | --- | --- | --- | --- |
| 3:98531235-TG-T | 71.09 | 99.83% | 32.04 | 99.93% |
| 3:98530074-T-A | 62.54 | 99.07% | 31.49 | 99.93% |
| 3:98541120-C-A | 52.13 | 99.84% | 32.7 | 100% |
| 3:98541153-A-T | 54.06 | 99.91%% | 33.36 | 100% |

**Supplemental Table 4** Results of gene-level collapsing analysis

(Table in a separate file)

**Supplemental Table 5** Clinical characteristics of 89 RP patients with and without *DCBLD2* ultra-rare damaging QVs

|  | RP with DCBLD2 ultra-rare damaging QVs | RP without DCBLD2 ultra-rare damaging QVs | P value |
| --- | --- | --- | --- |
| **Demographic Features** |  |  |  |
| Female sex | 74% | 76% | 1.00 |
| Age at the onset of symptoms (years, mean+-SD) | 23 +- 17 | 30 +- 16 | 0.24 |
| Age at diagnosis (years, mean+-SD) | 31 +- 20 | 37 +- 18 | 0.49 |
| **Clinical Manifestations** |  |  |  |
| Auricular chondritis | 86% | 61% | 0.25 |
| Nasal chondritis | 57% | 80% | 0.16 |
| Airway chondritis | 43% | 49% | 1.00 |
| Costochondritis | 57% | 74% | 0.38 |
| Ocular inflammation | 43% | 27% | 0.40 |
| Cutaneous inflammation | 50% | 23% | 0.17 |
| Arthritis or arthralgia | 86% | 87% | 1.00 |
| Oral ulcerations | 57% | 21% | 0.05 |
| Genital ulcerations | 29% | 15% | 0.30 |
| Venous thromboembolism^a^ | 14% | 5% | 0.34 |
| Raynaud’s phenomenon | 29% | 15% | 0.30 |
| Central nervous system involvement | 0% | 0% | 1.00 |
| Myocarditis | 0% | 0% | 1.00 |
| Pericarditis | 14% | 2% | 0.22 |
| Aortitis | 14% | 1% | 0.15 |
| Antinuclear antibody positive | 14% | 14% | 1.00 |
| **Treatment** |  |  |  |
| High-dose steroid^b^ | 57% | 58% | 1.00 |
| Immunosuppressants | 57% | 83% | 0.13 |
| Biological or targeted immunosuppressants | 43% | 70% | 0.21 |
| **Complications** |  |  |  |
| Death | 0% | 5% | 1.00 |
| ICU admission | 14% | 15% | 1.00 |
| Tracheotomy | 14% | 10% | 0.54 |
| Subglottic stenosis | 14% | 17% | 1.00 |
| Tracheomalacia | 29% | 41% | 0.70 |
| Bronchomalacia | 29% | 20% | 0.63 |

a. Venous thromboembolism includes unprovoked deep venous thrombosis and pulmonary embolism.
b. High-dose steroid means prednisone higher than 60 mg daily or equivalent.

**Supplemental Table 6** Clinical manifestations of patients with RP and DCBLD2 ultra-rare damaging QVs

(This table is removed given MedRxiv’s de-identification policy. Readers can contact the corresponding author for access to the data)

**Supplemental Table 7** Genes favorably weighted in the HALLMARK TNF pathway

(Table in a separate file)

**Supplemental Table 8** Pathway analysis using HC test

(Table in a separate file)

**Supplemental Table 9** Pathway analysis using GSEA

(Table in a separate file)

**Supplemental Table 10** Pathway Analysis using SKAT robust test

(Table in a separate file)

**Supplemental Table 11** Pathway analysis top hits

(Table in a separate file)

**Supplemental Figure 1** Flowchart of bioinformatic processing and statistical methods

(Table in a separate file)

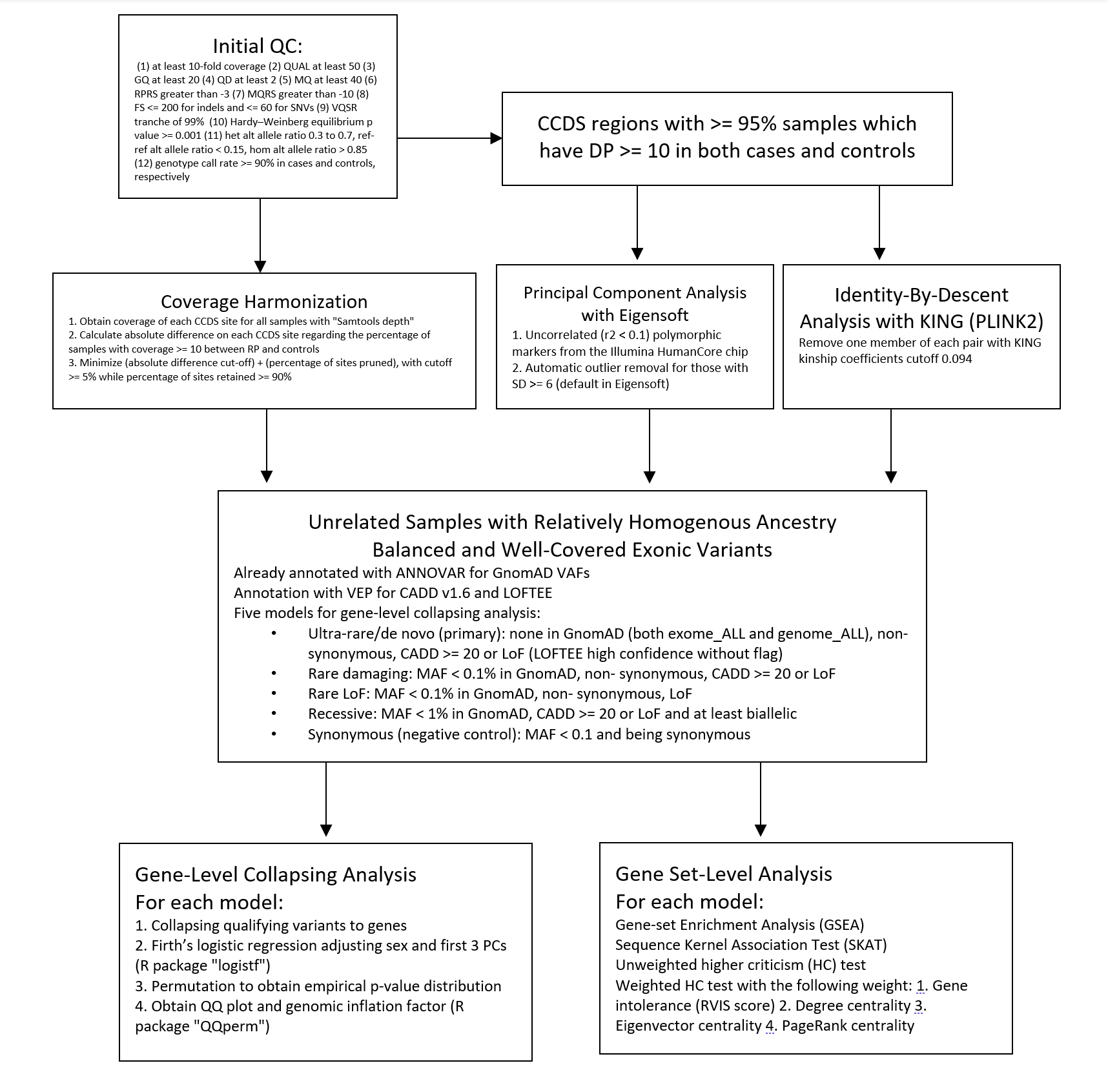

**Supplemental Figure 2** PCA plot of cases and controls

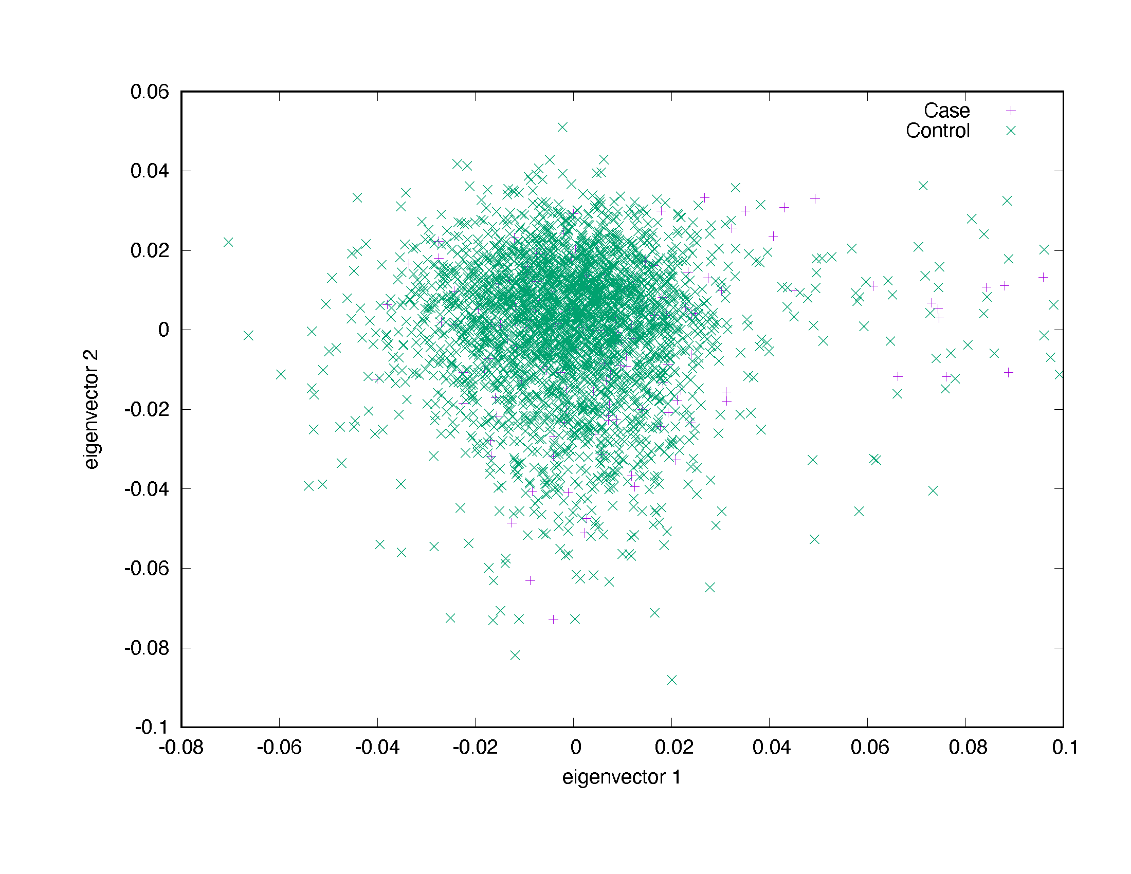

The PCA analysis examines population genetic structure and confirms that the cases and controls are from a homogenous ancestral cluster.

**Supplemental Figure 3** Amino acid position changes of the *DCBLD2* ultra-rare QVs and the affected domains

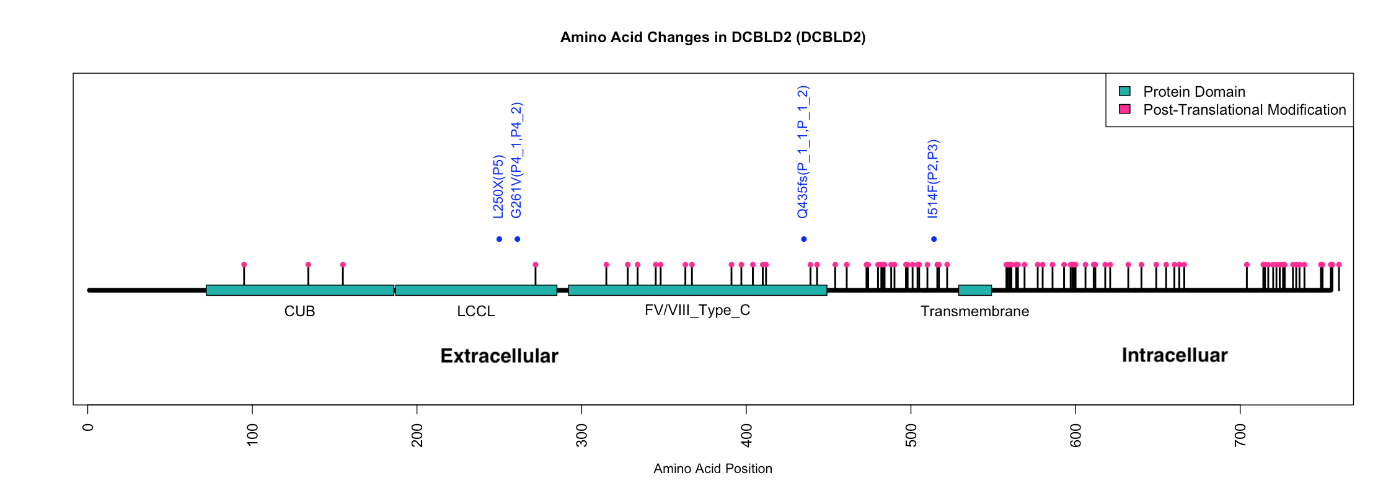

Each *DCBLD2* ultra-rare QV discovered in RP is located in the extracellular domain of DCBLD2

**Supplemental Figure 4** QQ plot of rare damaging model, PTV model, recessive model and synonymous model

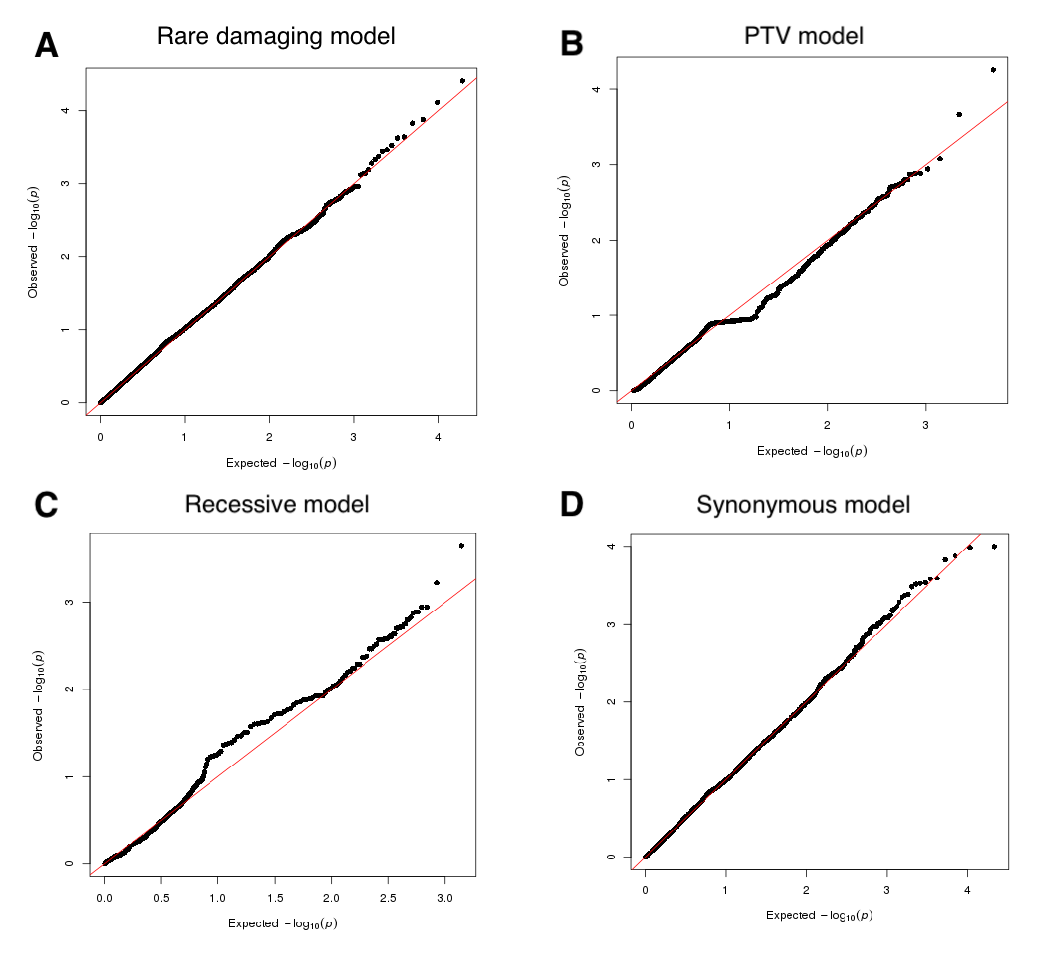

No genes were found associated with RP in the rare damaging model, PTV model, recessive model or synonymous model.

**Supplemental Figure 5** Sanger sequencing chromograms of the discovered *DCBLD2* ultra-rare damaging QVs

**Supplemental Figure 6** RELB protein interaction network

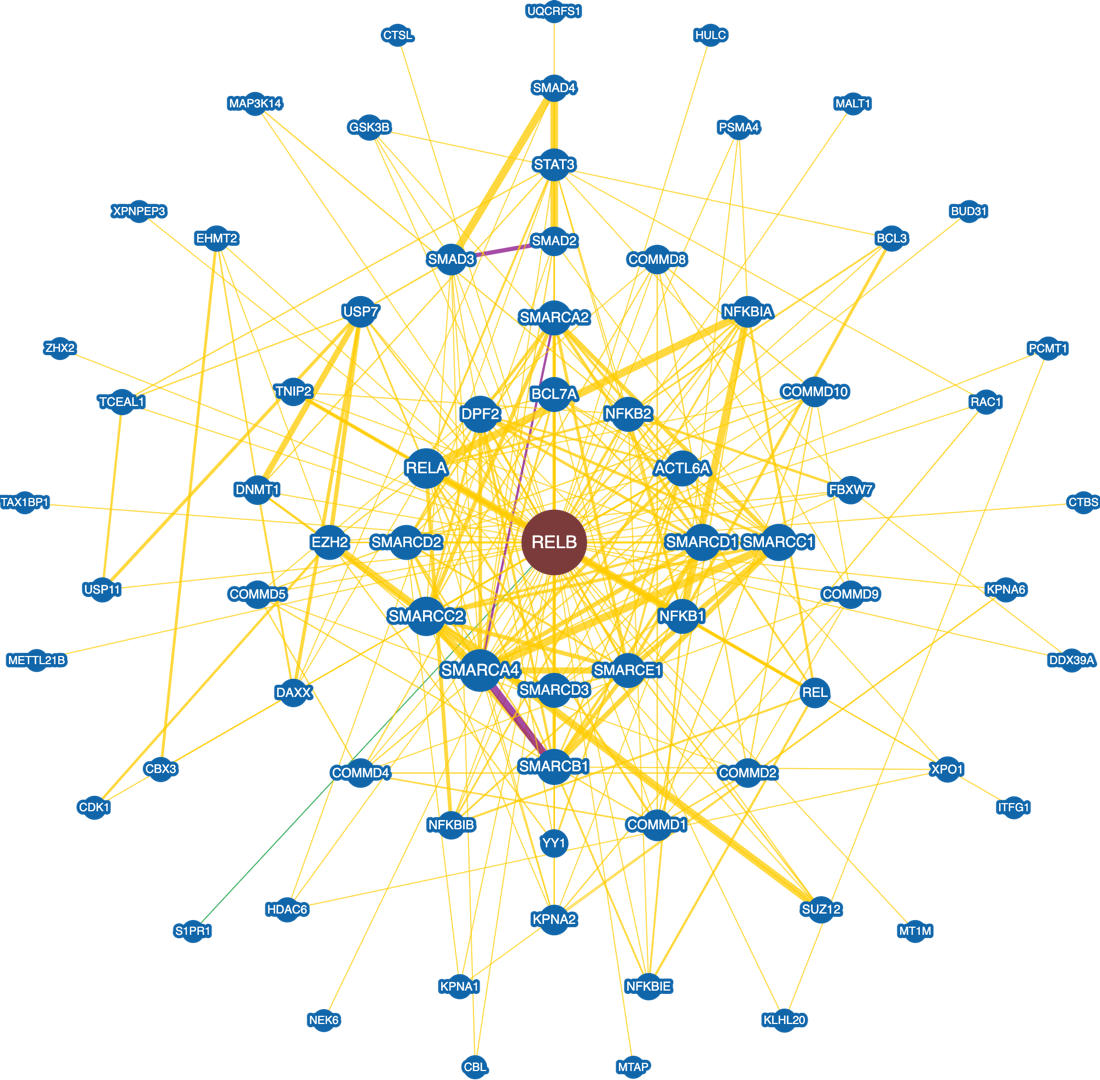

**Supplemental Figure 7** RELA protein interaction network

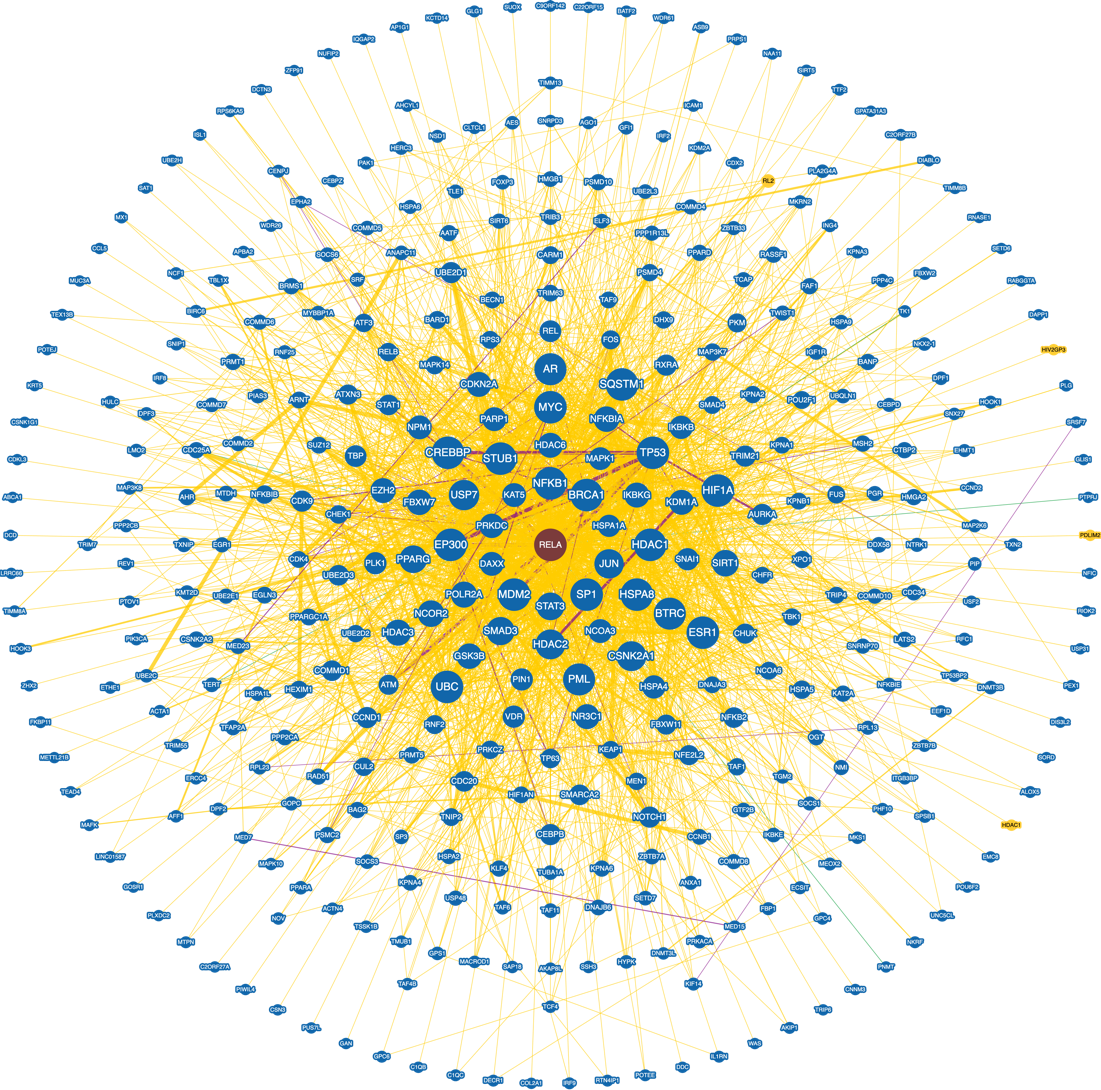

**Supplemental Figure 8** REL protein interaction network

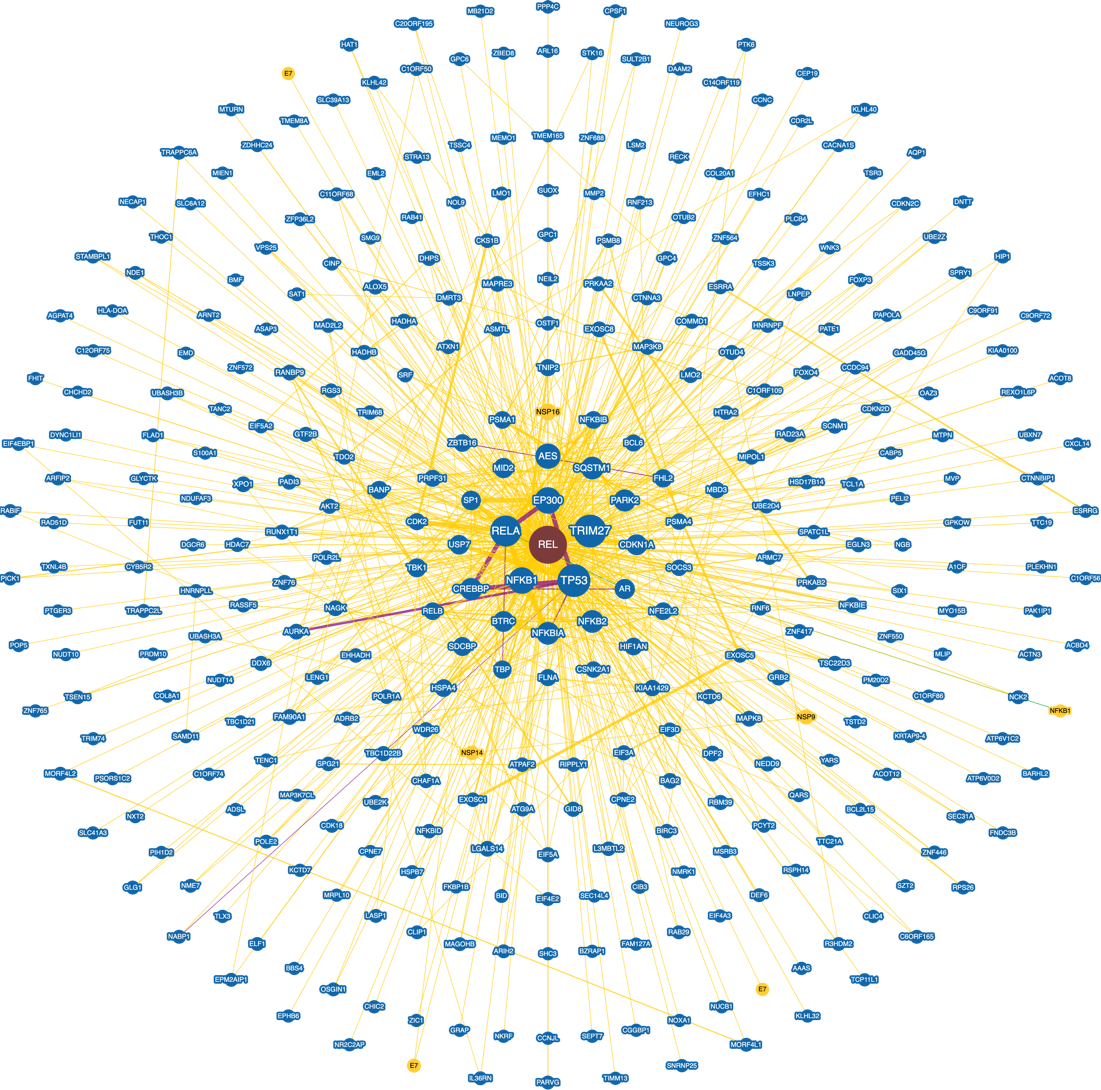
